# Cerebral aneurysm walls contain myoglobin that is possibly produced by myofibroblasts and contributes to wall thickening

**DOI:** 10.1101/2023.06.15.23291244

**Authors:** Hidehito Kimura, Tatsuya Mori, Kosuke Hayashi, Yusuke Ikeuchi, Kazuhiro Tanaka, Masakazu Shinohara, Akio Tomiyama, Eiji Kohmura, Takashi Sasayama

## Abstract

**Background:** Cerebral aneurysms are associated with subarachnoid hemorrhages if ruptured; however, mechanisms underlying aneurysmal wall thinning and thickening remain unclear.

We previously identified patterns of hemodynamic flow in aneurysms that are associated with wall thickening and thinning, and in this study our objective was to uncover the biological basis for these findings.

**Methods:** Cerebral aneurysmal wall samples were collected between August 2020 and March 2022 for proteomic analysis and immunohistological investigation of smooth muscle cells, myoglobin, and inducible nitric oxide synthase (iNOS) expression. We examined the co-localization of myoglobin expression within smooth muscle cells, identified by α-smooth muscle actin (α-SMA) staining, and myofibroblasts, identified by periostin staining. We measured collagen density in the samples using Sirius Red staining and investigated its correlation with myoglobin density.

**Results:** Analysis of proteins extracted from an area of thickening in the aneurysmal wall of one patient confirmed the presence of myoglobin. In 24 formalin-fixed aneurysmal wall samples, 19 expressed myoglobin, with 11 showing strong expression, and eight showing weak expression. Myoglobin was scattered or clustered within the vascular smooth muscle layer and tended to be expressed at sites other than where iNOS was identified. Double-label immunofluorescence staining confirmed that the myoglobin-positive rate within α-SMA-positive cells and α-SMA-positive areas was 33.2±23.8% and 31.3±37.8%, respectively, whereas within periostin-positive cells and periostin-positive areas it was 92.2±13.7% and 79.8±29.5, respectively. A moderate correlation was observed between the density of myoglobin and collagen in the same sample field, with a Spearman’s rank correlation coefficient of 0.593 (p = 0.036).

**Conclusions:** Cerebral aneurysmal walls express myoglobin, which may be produced by myofibroblasts in the wall. Areas with high myoglobin levels retain high levels of collagen fibers, and myoglobin may be involved in wall thickening by suppressing destructive changes in the extracellular matrix collagen fibers.

## Introduction

Recent advances in imaging technology have increased the frequency (about 1–5% of the general population) with which unruptured cerebral aneurysms can be identified.^1–3^ Cerebral aneurysms can cause subarachnoid hemorrhages if ruptured, which can be fatal.^4^ Most unruptured aneurysms are asymptomatic and do not change in shape; however, some aneurysms are known to rupture after enlargement.^5^ It is recommended to identify aneurysms that display a higher likelihood of rupture and prevent rupture with surgical intervention.^6^

Previous studies have predicted the characteristics of aneurysms that are prone to rupture based on factors such as size, location, shape, and patient background. Although larger aneurysms are more likely to rupture, small cerebral aneurysms can still rupture.^7–10^ The pathophysiology of aneurysm rupture remains unclear; therefore, it is difficult to predict the growth or rupture of individual aneurysms.

Recent advances in histopathological and molecular biology studies of cerebral aneurysms have shed light on aneurysmal formation, enlargement, and rupture.^11, 12^ A cerebral aneurysm is a localized bulge of a cerebral blood vessel pathologically characterized by degenerative changes in the wall. Aneurysmal growth is considered to be induced by mechanical damage to the vascular endothelium from hemodynamic stress on the vessel wall, that is, increased wall shear stress, which leads to the disappearance of the internal elastic lamina and degeneration of the smooth muscle tunica media layer. The vulnerable areas become aneurysms.^13^ Vulnerability is believed to be caused by chronic inflammation, mainly involving macrophages.^14^ Activated macrophages and smooth muscle cells (SMCs) produce macrophage-derived matrix metalloproteinase (MMP)-2, MMP-9, inducible nitric oxide synthase (iNOS), and reactive oxygen species (ROS), all of which induce extracellular matrix (ECM) degeneration, and SMC apoptosis, thereby promoting aneurysm formation. ^6, 13, 15^ As the ECM in the tunica media degenerates, the tunica media thins, and eventually only the tunica externa remains.^16^

These observations explain why the aneurysmal walls become fragile and thin, and since aneurysms are believed to rupture at the thinning part of the wall, they clarify the steps leading to aneurysm rupture.^12^ However, this does not explain why thick- and thin-walled regions coexist within a single aneurysm. Uncovering the precise mechanisms responsible for the disparity between the thin- and thick-walled regions of the aneurysmal wall is crucial for elucidating the true mechanism of rupture.

We previously performed a computational fluid dynamics (CFD) analysis of cerebral aneurysms to elucidate the hemodynamic factors that induce wall thinning and thickening. We found that the degree of blood flow stagnation is related to the degree of wall thinning and thickening. In other words, we found that the aneurysmal wall is more likely to be thinned in areas where the wall shear stress vector hardly fluctuates and the blood flow is unidirectional during one cardiac cycle, whereas the wall is more likely to be thickened in areas where the wall shear stress vector dramatically fluctuates and the flow direction tends to change in multiple directions.^17, 18^ These findings were consistent with another published report.^19^

To elucidate the biological mechanisms by which these hemodynamic differences induce thickness change of the aneurysmal wall, we performed a proteome analysis to detect differentially expressed proteins in the thin or thick portions of the aneurysmal wall and compared protein expression levels and location by immunohistochemistry.

## Methods

### Patients

This study was conducted in accordance with criteria from the Strengthening the Reporting of Observational Study in Epidemiology statement and approved by the local ethics committee of our institution (#B210170).^20^ Written informed consent was obtained from all the patients. Patients who underwent clipping of unruptured cerebral aneurysms at our hospital between August 2020 and March 2022 were included in this study. The aneurysms were sampled after clipping to occlude their blood flow. If the aneurysm was deep-seated or small, then excision of the aneurysmal wall was not performed because of technical difficulty. The aneurysm was observed under an operating microscope before clipping; thin-walled areas appeared red, and thickened areas of the wall were uniformly white or white-yellow. Samples were collected from the wall, as widely separated as possible. The samples were stored frozen or infiltrated with formalin. A total of 29 specimens were successfully obtained from 21 patients who underwent aneurysmal clipping. There were 10 men and 11 women, and their mean age was 65.5±13.1 years. There were 10 middle cerebral artery, six anterior cerebral artery, and five internal cerebral artery aneurysms; 16 aneurysms were unruptured and five had ruptured. Mean aneurysm seize was 6.68 mm (range: 4.5–10.1 mm).

A total of 11 frozen specimens of arterial wall (four thickened, seven thinned) were obtained from nine out of 12 patients, including two patients from whom samples from both thinned and thickened areas of the wall were obtained. Samples were not collected from the remaining three patients because of technical difficulties. Proteome analysis was performed on these 11 specimens. A further 24 formalin-fixed samples (10 thickened, six thinned, and eight mixed-wall regions) were collected from 17 patients for immunohistochemical analysis (Suppl Fig. 1).

### Proteome analysis

(1) Protein extraction: The required volume of lysis buffer was prepared according to the procedure of the protein extraction kit (Nuclear Extract Kit; Active Motif, Carlsbad, CA, USA), the sample was added, and tissue disruption was performed using a sonicator. The amount of protein in the extract was measured using a protein quantification reagent (Pierce BCA Protein Assay Kit; Thermo Fisher Scientific, Waltham, MA, USA), and the samples were prepared for electrophoresis.
(2) Protein electrophoresis and staining: Samples were electrophoresed on sodium dodecyl-sulphate (SDS)-polyacrylamide gels. Silver staining was performed after electrophoresis using a silver staining kit (ProteoSilver Plus Silver Stain Kit; Sigma-Aldrich) according to the kit instructions.
(3) Liquid chromatography-tandem mass spectrometry (LC-MS/MS). Silver-stained protein bands that were specifically observed in either the thickened or thinned aneurysmal wall areas were identified and excised. LC-MS/MS analysis was performed using an LCMS-IT-TOF instrument (Shimadzu, Kyoto, Japan) interfaced with a nanoreverse-phase liquid chromatography system (Shimadzu). MS/MS data were analyzed using Mascot (version 2.3.01; Matrix Science, London, UK), as previously reported.^21^ The search parameters were as follows: enzyme, trypsin; variable modifications, carbamidomethyl (Cys), deamidated (Asn, Gln), and oxidation (Met); peptide mass tolerance, ±50 ppm; fragment mass tolerance,±0.05 Da; and max missed cleavages.^21^ Western blot analysis

Western blotting was performed to confirm the myoglobin expression. We used the following antibody: anti-myoglobin (diluted 1:500, SC-74525; Santa Cruz Biotechnology, Dallas, TX, USA).

### Immunohistological analysis

Immunohistochemical analysis was performed to detect myoglobin expression in the aneurysmal wall, to observe the characteristics of the expression site and localization, and the relationship between SMCs and iNOS expression. The specimens were fixed in 4% formaldehyde and embedded in paraffin. The samples were originally 2–3 mm in size, and 2-μm-thick axial slices were prepared. The sections were deparaffinized and immersed in methanol containing 3% hydrogen peroxide. Antigen retrieval was performed by heat mediation in a 0.01 mol/L citrate buffer (pH 6.0) for 15 min by autoclaving (121 °C, 2 atm). The sections were then incubated with primary antibodies against α-smooth muscle actin (α-SMA; diluted 1: 100, ab5694; Abcam, Cambridge, UK), myoglobin (diluted 1: 500, SC-74525, Santa Cruz Biotechnology, Dallas, TX), and iNOS (diluted 1:1000, 18985-1-AP, Proteintech group, Manchester, UK) at 4 °C overnight. The sections were allowed to react with a peroxidase-conjugated anti-rabbit IgG polyclonal antibody or anti-mouse IgG polyclonal antibody (Histofine Simple Stain MAX-PO; Nichirei, Tokyo, Japan) for 60 min, and the reaction products were visualized by immersing the sections in 0.03% diaminobenzidine solution containing 2 mM hydrogen peroxide for 1–5 min.

Double-label immunofluorescence staining was used to investigate the origin of myoglobin in aneurysmal walls. α-SMA-positive cells have been reported to include SMCs and myofibroblasts, which are also known to have myoglobin. Therefore, we searched for the presence of myofibroblasts in the specimens.^22–24^ Periostin was selected as a biomarker of myofibroblasts.^22, 25, 26^ Staining was performed using the ImmPRESS Duet Double Staining Polymer Kit (Vector Laboratories, Burlingame, CA, USA). The sections were incubated with mouse primary antibodies against α-SMA (dilution 1:100, ab5694, Abcam, United Kingdom), myoglobin (diluted 1:500, SC-74525, Santa Cruz Biotechnology, Dallas, TX, USA), and rabbit primary antibodies against periostin (dilution 1:400, Proteintech, 19899-1-AP) for 1 h at room temperature.

The sections were then incubated with VectorFluor Duet Reagent for 30 min. Coverslips were mounted using ProLongTM Gold Antifade Reagent with DAPI (Life Technologies). The images were acquired using a Keyence BZ-X700 fluorescence microscope. The numbers of immunopositive cells and immunopositive areas were measured in the entire field of the photographs. The results are presented as the average value for each patient (cells/field). The colocalization ratio was calculated and represented as a percentage using a BZ-X analyzer.

In addition, Sirius Red staining was performed to quantify the density of collagen. Uniformly concentrated areas on the Sirius Red-stained photomicrograph were randomly selected (200–300 μm^2^), and the same areas of the fluorescent myoglobin-stained photomicrographs were marked. The percentages of collagen- and myoglobin-containing areas in the selected fields were quantified using image analysis software (Image J 1.48v, National Institutes of Health), and correlations were examined.^27^

### Statistical analysis

All continuous quantitative data were represented as means ± standard deviation (SD) and categorical variables as percentages. The Mann–Whitney U test was used to evaluate the co-localization rate of myoglobin with α-SMA and periostin, and periostin with α-SMA. Spearman’s rank correlation test was performed to examine potential correlations between the Sirius red-positive areas and myoglobin-positive areas. The degree of association was interpreted as follows: strong (absolute value of correlation coefficient, |*r*| = 0.7–1), moderate (|*r|* = 0.5–0.7), or low (|*r*| = 0.3–0.5).

All statistical analyses were performed using EZR (Saitama Medical Center, Jichi Medical University, Saitama, Japan), which is a graphical user interface for R 3.4.3 (R Foundation for Statistical Computing, Vienna, Austria). A two-sided p < 0.05 was considered statistically significant.

## Results

### Detection of myoglobin

Silver staining was performed on the protein electrophoresis gels of samples from 11 aneurysmal wall sites (four thickened and seven thinned walls), including two thin and thick paired regions. In one of the two patients with samples from both thinned and thickened wall regions, we observed a prominent band that appeared only in the sample from the thickened area of the wall (Fig. 1). The equivalent band was faint in the protein electrophoresis gel from the second patient with both thick and thin wall areas, and was not clearly seen in samples from the remaining two thickened sites. Hence, this band was detected intensely in one of the four samples from the thickened areas. This band was extracted and subjected to proteomic analysis, which identified it as human myoglobin protein.

**Figure 1.**
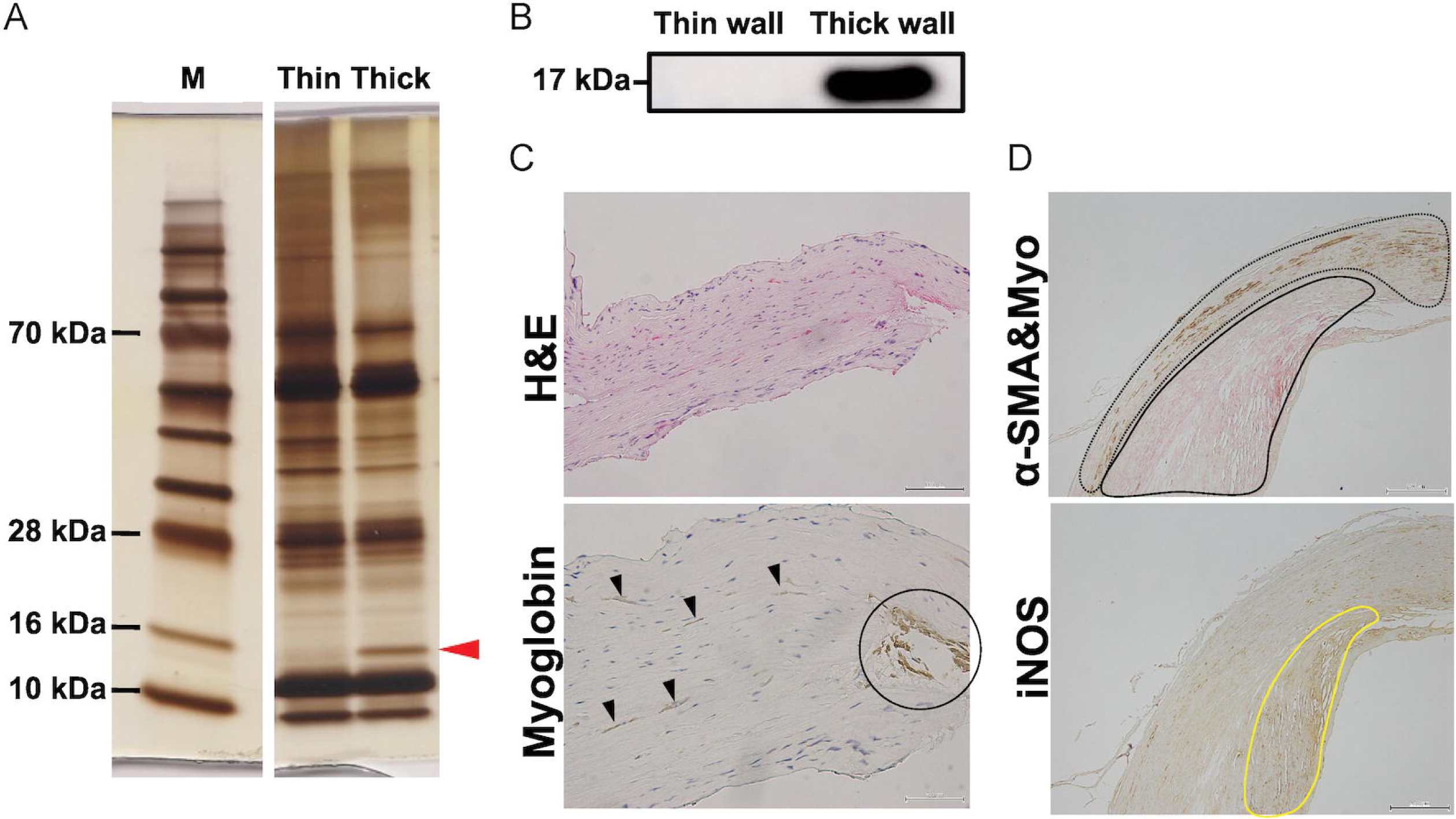
Detection of human myoglobin protein and its expression in the aneurysmal wall A. Silver-stained sodium dodecyl sulphate-polyacrylamide gel electrophoresis of proteins extracted from thinned and thickened aneurysmal walls. A prominent band was detected in the sample from the thickened wall area (arrowhead). M: molecular weight marker. B. Western blot analysis of the excised band using a primary antibody against myoglobin. C. Aneurysmal wall (H&E, and myoglobin immunohistochemical staining). Myoglobin was found scattered (arrowheads) in the media, or clumped in other areas (encircled). D. Immunohistological examination (upper: double staining for myoglobin (pink) and α-SMA (brown); lower: single staining for iNOS) revealed that myoglobin was expressed inside (encircled by the black line in the upper photograph) and surrounding the iNOS expression area (encircled by the yellow line in the lower photograph), and α-SMA-positive vascular smooth muscle was present outside this site (encircled by the black dotted line in the upper photograph). α-SMA: α-smooth muscle actin; H&E: hematoxylin and eosin; iNOS: inducible nitric oxide synthase

Western blot analysis confirmed the presence of myoglobin in the thickened walls (Fig. 1).

### Location and distribution of myoglobin in the aneurysmal wall

Among the 24 formalin-fixed samples (10 thickened, six thinned, and eight mixed-wall regions), 19 expressed myoglobin, with strong expression in 11 and weak expression in remaining eight. For the six specimens from thinned wall regions, two had faint myoglobin expression, and the remaining four showed no expression. SMC and iNOS were found in all the samples. Regarding the distribution of myoglobin, some areas were scattered and dispersed within the SMC layer of the wall, while others were relatively clustered in the vicinity of the SMC layer (Fig. 2).

**Figure 2.**
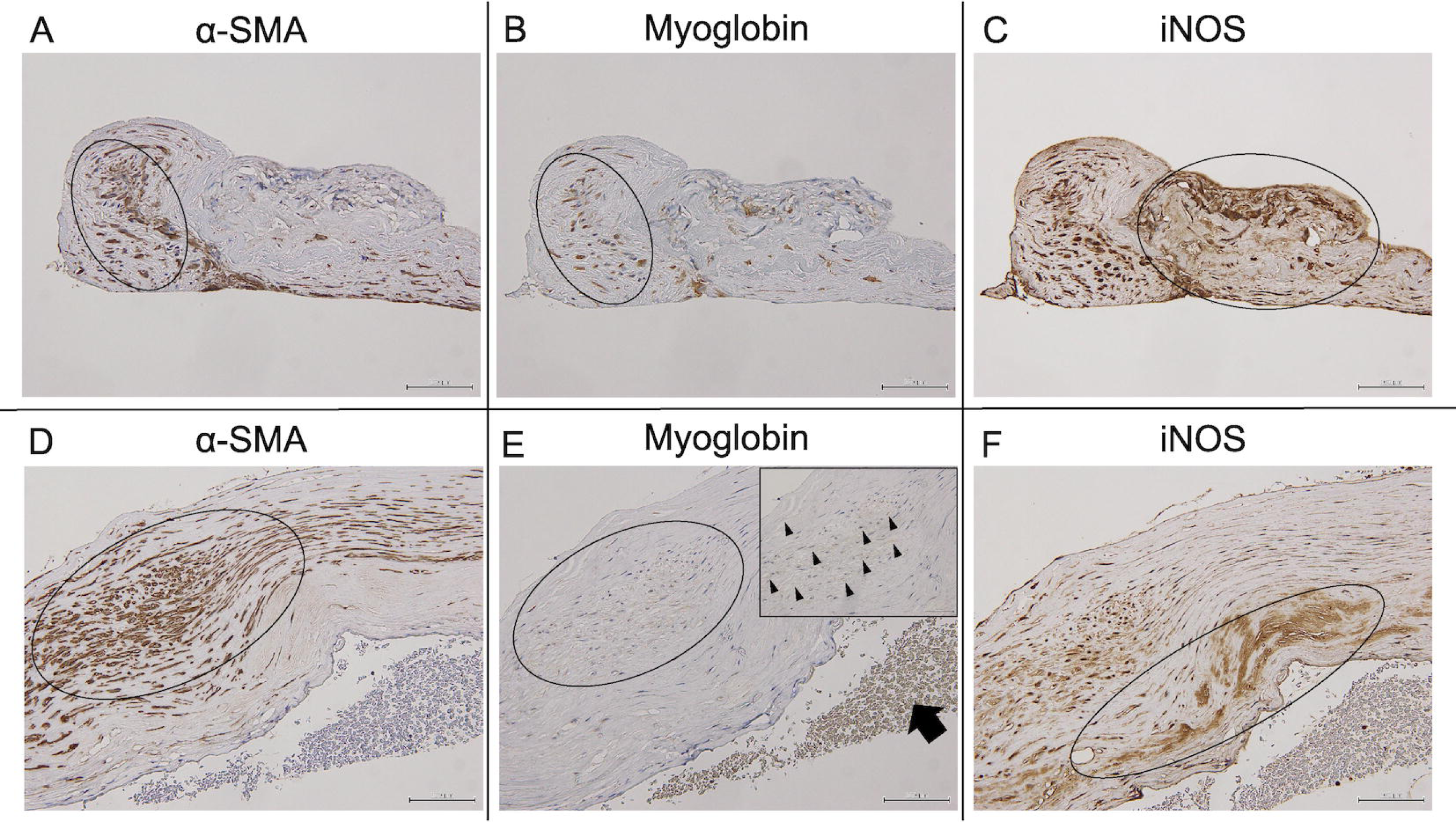
Expression of α-SMA, myoglobin and iNOS in the aneurysmal wall Photomicrographs of immunohistochemistry using primary antibodies against α-SMA (alpha-smooth muscle actin; left), myoglobin (middle), or iNOS (inducible nitric oxide synthase; right) in a representative case with strong and weak expression of myoglobin (A–C, D–F; respectively). Myoglobin was found scattered (encircled in B, E; arrowhead in the inset, magnified view of the encircled area in E) throughout the smooth muscle cell cluster (defined by the presence of α-SMA, encircled in A, D), but also clumped in other areas (arrow in E). iNOS tended to be expressed at the sites where α-SMA was not (encircled areas in C, F). Scale bar = 100 μm

In contrast, iNOS-expressing regions were generally more sparse and with coarse staining, and myoglobin expression was less likely to occur in these regions. On the other hand, myoglobin-expressing regions were relatively densely packed within the SMC layers.

### Origin of myoglobin

Double-fluorescence immunohistological examination revealed scattered myoglobin in the regions containing α-SMA, but also in other areas without α-SMA (Fig. 3). The co-localization rate of myoglobin within α-SMA positive cells was 33.2±23.8%, and 31.3±37.8% in the α-SMA-containing area. On the other hand, the co-localization rate of myoglobin within periostin-positive cells was 92.2±13.7%, and 79.8±29.5 in the periostin-containing area; the distribution of periostin and myoglobin was almost identical (Fig. 3).

**Figure 3.**
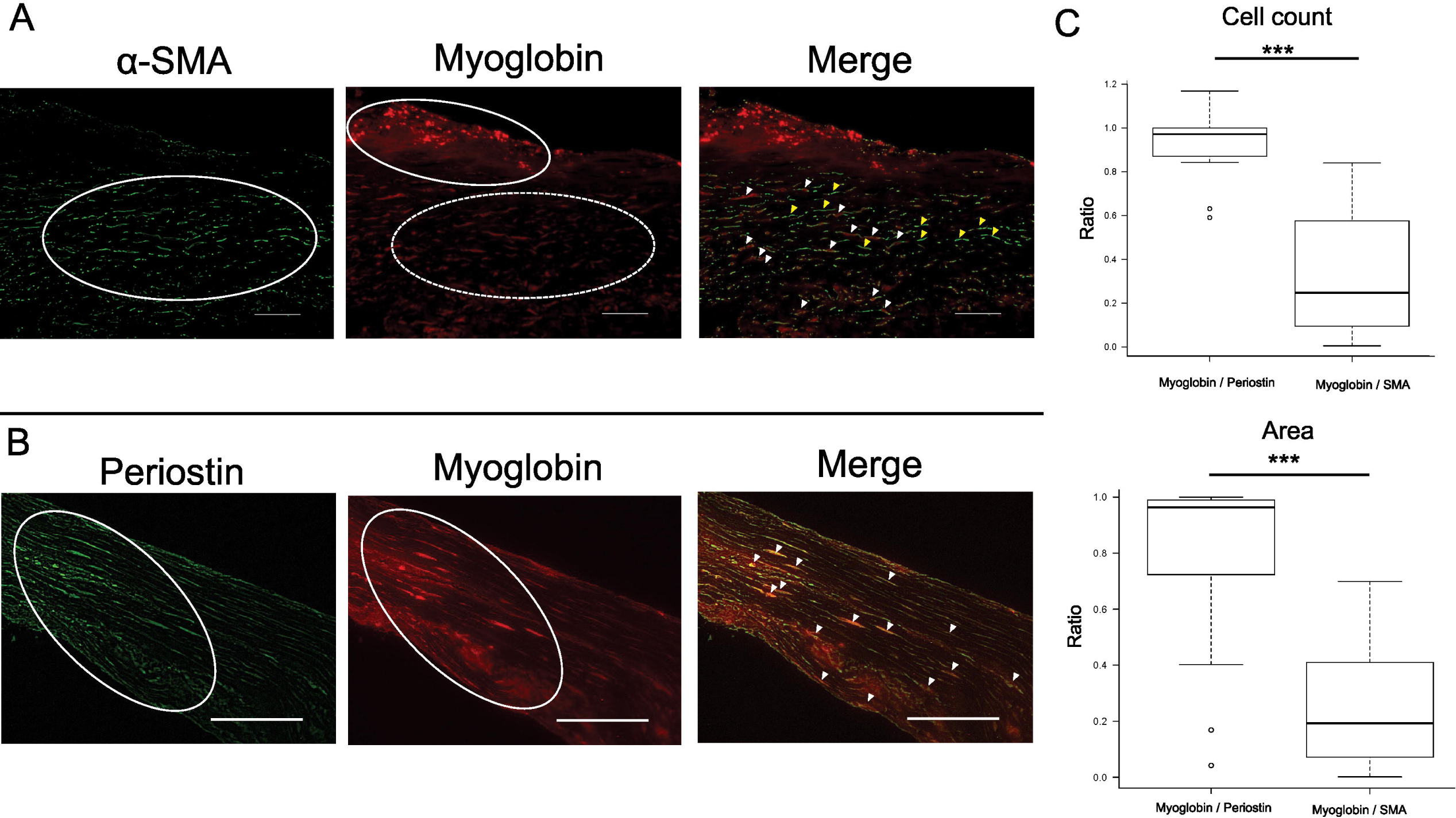
Expression of myoglobin in α-SMA- and periostin-positive cells and areas Photomicrographs of double immunofluorescence using primary antibodies against α-SMA (alpha-smooth muscle actin; A, left) and myoglobin (A, middle) and the merged image (A, right) in a section from an aneurysm, and periostin (B, left) myoglobin (B, middle) and the merged image (B, right) in a section from another aneurysm.The main distribution of α-SMA-positive cells (encircled in A left) is inconsistent with the distribution of myoglobin- positive cell clusters (encircled in A middle). Cells showing weak myoglobin staining were scattered in the smooth muscle layer (dotted circles in A middle).The merged image of α-SMA and myoglobin (A right) showed that some α-SMA-positive cells expressed myoglobin (white arrowheads as representative, A right), but others did not (yellow arrowheads as representative, A right). In contrast, the distribution of periostin-positive cells (encircled in B left) was almost consistent with the distribution of myoglobin (encircled in B middle). The merged image demonstrates that almost all periostin-positive cells also expressed myoglobin (arrowheads as representative, B right). The difference of co-localization ratio of myoglobin within α-SMA-positive cells and within periostin-positive cells (C, upper) was statistically significant (*** p <0.001) and the difference of co-localization ratio of myoglobin within α-SMA-positive areas and within periostin-positive areas (C, lower) was also statistically significant (^IZIZIZ^ p <0.001) . Scale bar = 100 μm

The difference of co-localization ratios of myoglobin within α-SMA-positive cells and within periostin-positive cells (Fig. 3C, upper panel) was statistically significant (p<0.001) and difference of co-localization ratios of myoglobin within α-SMA-positive areas and within periostin-positive areas (Fig. 3C, lower panel) was also statistically significant (p<0.001). Receiver operating characteristic curve analysis showed that there were no significant difference between comparison of within-cell co-localization (myoglobin/periostin, vs myoglobin/α-SMA, solid line in Suppl Fig. 2; area under the curve (AUC) 0.975 [95% CI 0.937 – 1]) and area co-localization (myoglobin/periostin vs myoglobin/α-SMA, dashed line in Suppl Fig. 2; AUC 0.867 [95% CI 0.734 - 0.999]), p = 0.0525.

After the double staining of α-SMA and periostin, the co-localization rate of periostin within α-SMA-positive areas was 41.8±24.3%, and 57.5±24.8% in the α-SMA-positive cells (Fig. 4).

**Figure 4.**
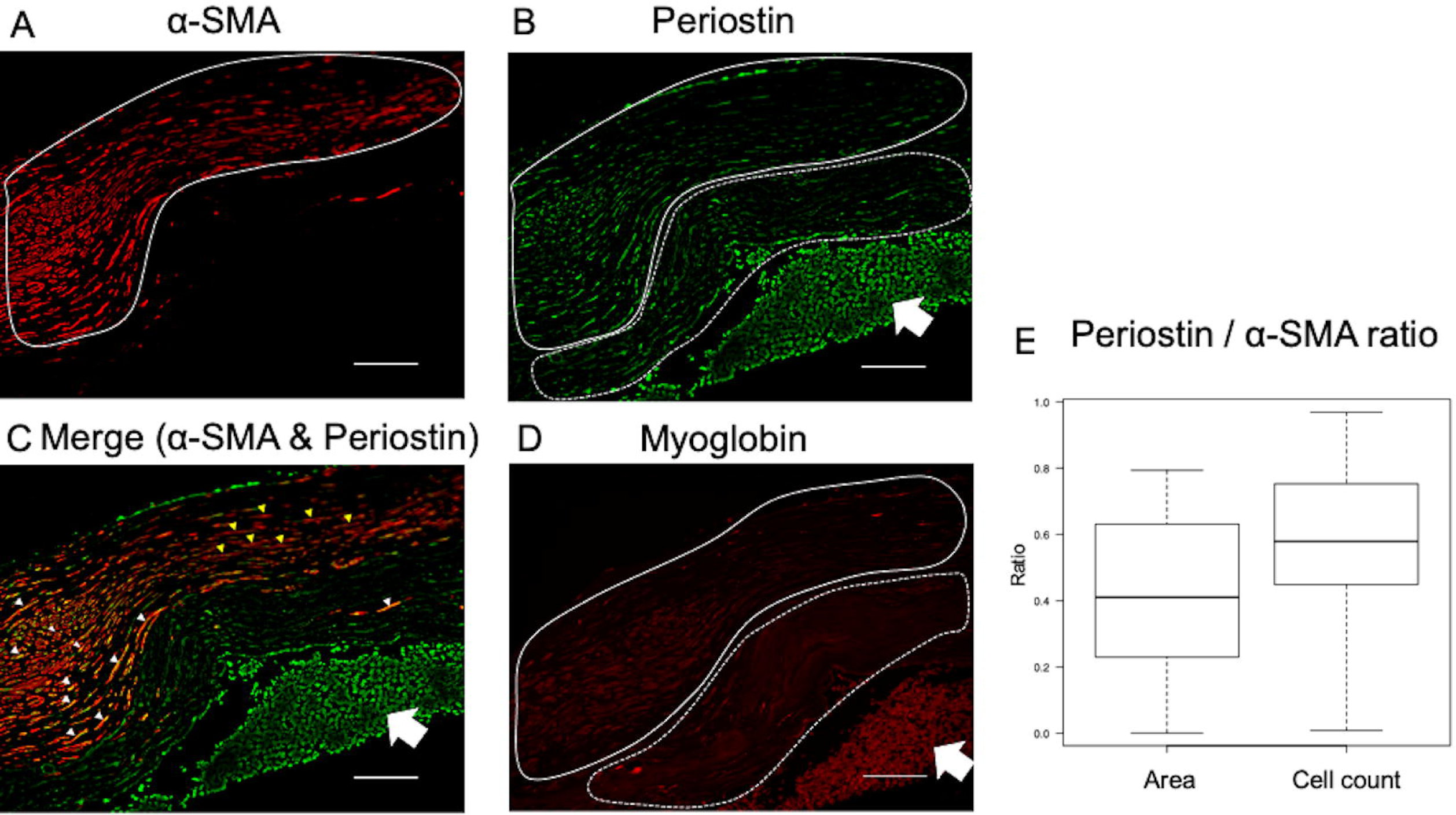
Relationship of α-SMA, periostin and myoglobin in the aneurysmal wall. Photomicrographs of double immunofluorescence using primary antibodies against (A) α-SMA (alpha-smooth muscle actin), periostin (B), with merged image of α-SMA and periostin (C), and myoglobin (D) (The sections are from the same patient as in Figure 2 lower row.) The α-SMA-positive cell clusters (delineated by the white line in A) are partially overlapped with the periostin-positive cells (delineated by white line in B). In contrast, outside of α-SMA-positive cell clusters, other periostin-positive cells were also found (delineated by dotted line in B). The merged image of α-SMA and periostin (C) showed that some α-SMA postive-cells expressed periostin (white arrowheads as representative), but others did not or had very weak expression (yellow arrowheads as representative). Myoglobin-positive cells (D) were expressed in areas consistent with periostin-positive cells, delineated both with solid and dotted line, and corresponding areas in B, including strong periostin-positive cluster areas (arrow), as seen in B, C and Figure 2 E. The co-localization rate of periostin within α-SMA-positive areas was 41.8±24.3%, and 57.5±24.8% in the α-SMA-positive cells (E). Scale bar = 100 μm

### Relationship between myoglobin density and collagen fiber density

On a photomicrograph of a Sirius Red-stained section of an aneurysm wall, a uniform area of tissue was selected for measurement of collagen density. The same area of tissue section was evaluated for myoglobin density on a photomicrograph of a myoglobin-stained section. Fourteen areas from 12 samples were compared. A moderate correlation was observed, with a Spearman’s rank correlation coefficient of 0.593 (p = 0.036) (Fig. 5).

**Figure 5.**
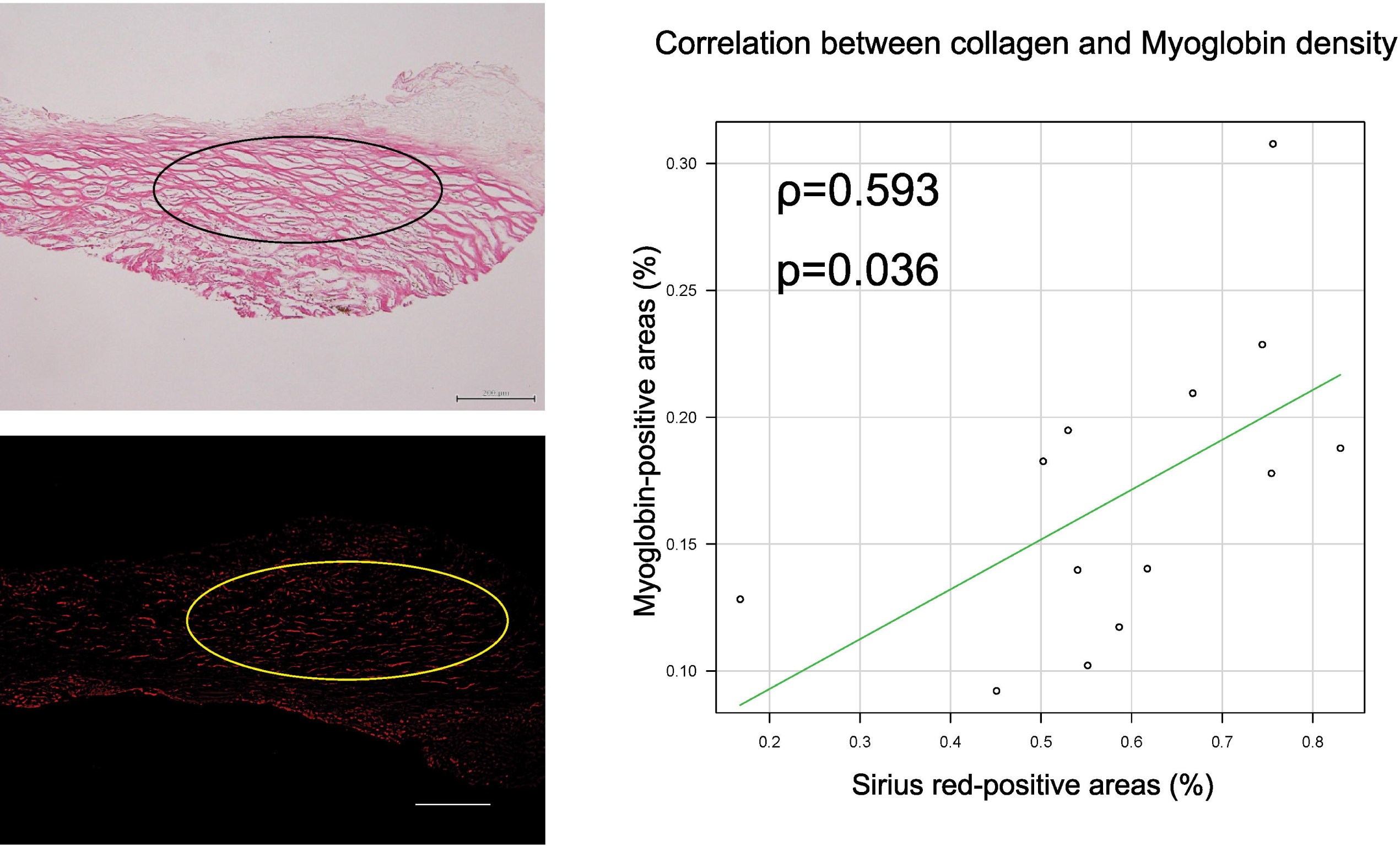
Correlation between Sirius Red-positive collagen and myoglobin density. In the Sirius Red-stained image (top left), uniform Sirius Red-positive areas were selected (circled), and the percentage of the Sirius Red-positive area was calculated (45.0% in this example). The equivalent area on the myoglobin-stained fluorescence photograph (lower left) was selected and the percentage of the myoglobin-positive area was calculated (0.09% in this example). Spearman’s rank correlation coefficient was analyzed by plotting data from 14 regions of 12 samples. A correlation was found between the two densities (ρ = 0.593, p = 0.036). Scale bar = 200 μm.

## Discussion

To the best of our knowledge, this is the first report of myoglobin detection in the human cerebral aneurysmal wall. Furthermore, we identified periostin as a potential indicator of myofibroblasts within the aneurysmal wall where myoglobin is present.

Here, we discuss the characteristics, possible origin, and role of myoglobin in aneurysmal walls.

### Characteristics of myoglobin expression in the aneurysmal wall

Myoglobin is a well-known cytoplasmic heme protein that is found in cardiac and skeletal muscle cells as an oxygen storage protein. Myoglobin is present in vascular smooth muscle, however, no reports have shown that myoglobin is expressed in cerebral aneurysm walls.^28^ In this study, myoglobin was mostly observed in the thickened areas of the aneurysmal wall, and less abundantly in the thinned areas. Myoglobin localization differed from that of iNOS, which was more frequently expressed in areas with scarce myoglobin (Fig. 2).

Myoglobin was found to be scattered between and inside SMCs in some areas of the tunica media, and clustered in the vicinity of the SMC layer in other areas (Fig. 1 C, D, Fig. 2). Myoglobin was commonly located between the smooth muscle layer and regions that exhibited elevated iNOS expression, suggesting its presence acted as a barrier to iNOS infiltration (Fig. 1 D).

### Origin of myoglobin in aneurysms

Myoglobin is reported to be expressed in the vascular SMC layer; therefore, we initially assumed that myoglobin in the aneurysmal wall was derived from vascular smooth muscle cells.^29^ Consequently, we performed double staining for α-SMA and myoglobin, but found that the expression sites only partially overlapped; the myoglobin-positive rate within α-SMA-positive cells was 33.2±23.8%, and within α-SMA-positive areas was 31.3±37.8%. This means only approximately 30% of the α-SMA-positive cells showed expression of myoglobin, and approximately 70% did not.

α-SMA-positive cells are known to include vascular smooth muscle and myofibroblasts.^22^ In addition, myofibroblasts have been recently reported to produce myoglobin.^25, 26^

We hypothesized that myofibroblasts exist in the aneurysmal wall and produce myoglobin. Hence, we performed immunofluorescence staining using periostin, a specific marker for myofibroblasts.^22, 25, 26^ Periostin expression was confirmed in all thickened areas of the aneurysmal wall. The myoglobin-positive rates within the periostin-positive cells and periostin-positive areas were 92.2±13.7% and 79.8±29.5, respectively, indicating that myoglobin was expressed in about 80–90% of the periostin-positive cells and areas. This demonstrates that myofibroblasts certainly exist in the aneurysmal wall and that many of them express myoglobin.

As regards the accuracy of differentiating myoglobin-positive rates in periostin-positive cells and areas compared with SMA-positive cells and areas, the cell count co-localization approach demonstrated marginally better accuracy, however the difference was not statistically significant (p = 0.0525 Suppl Fig. 2).

Furthermore, the periostin-positive rates within all α-SMA-positive areas and α-SMA-positive cells were 41.8±24.3% and 57.5±24.8%, respectively, suggesting that about half of the cells previously thought to be vascular smooth muscle may have been myofibroblasts.

### Significance of myoglobin in the aneurysmal wall

Myoglobin has been reported to function as a scavenger of NO and ROS.^30, 31^ NO is produced by iNOS within inflammatory cells, generates toxic proteins that impair the function of cytoskeletal proteins involved in ECM remodeling, and causes aneurysmal development.^32, 33^ ROS are produced by macrophages, are major mediators of various inflammatory cascades, and are known to promote aneurysmal development as well.^12, 34^ These macrophage-derived proteinases promote the degradation of the ECM and decrease collagen production at the aneurysmal wall, leading to thinning of the media and subsequent weakening/rupture.^35^

In this study, we found that the density of myoglobin in the cerebral aneurysmal walls correlated with the density of collagen fibers. Myoglobin is suggested to inactivate these macrophage-derived proteinases and suppress the degradation of the ECM, particularly collagen, in the aneurysmal wall. The myoglobin found in the aneurysmal walls in this study was found mostly in the thickened areas and less in the thinned areas, suggesting that myoglobin may prevent wall thinning. Figure 6 presents a potential biological role of myoglobin in aneurysmal walls.

**Figure 6.**
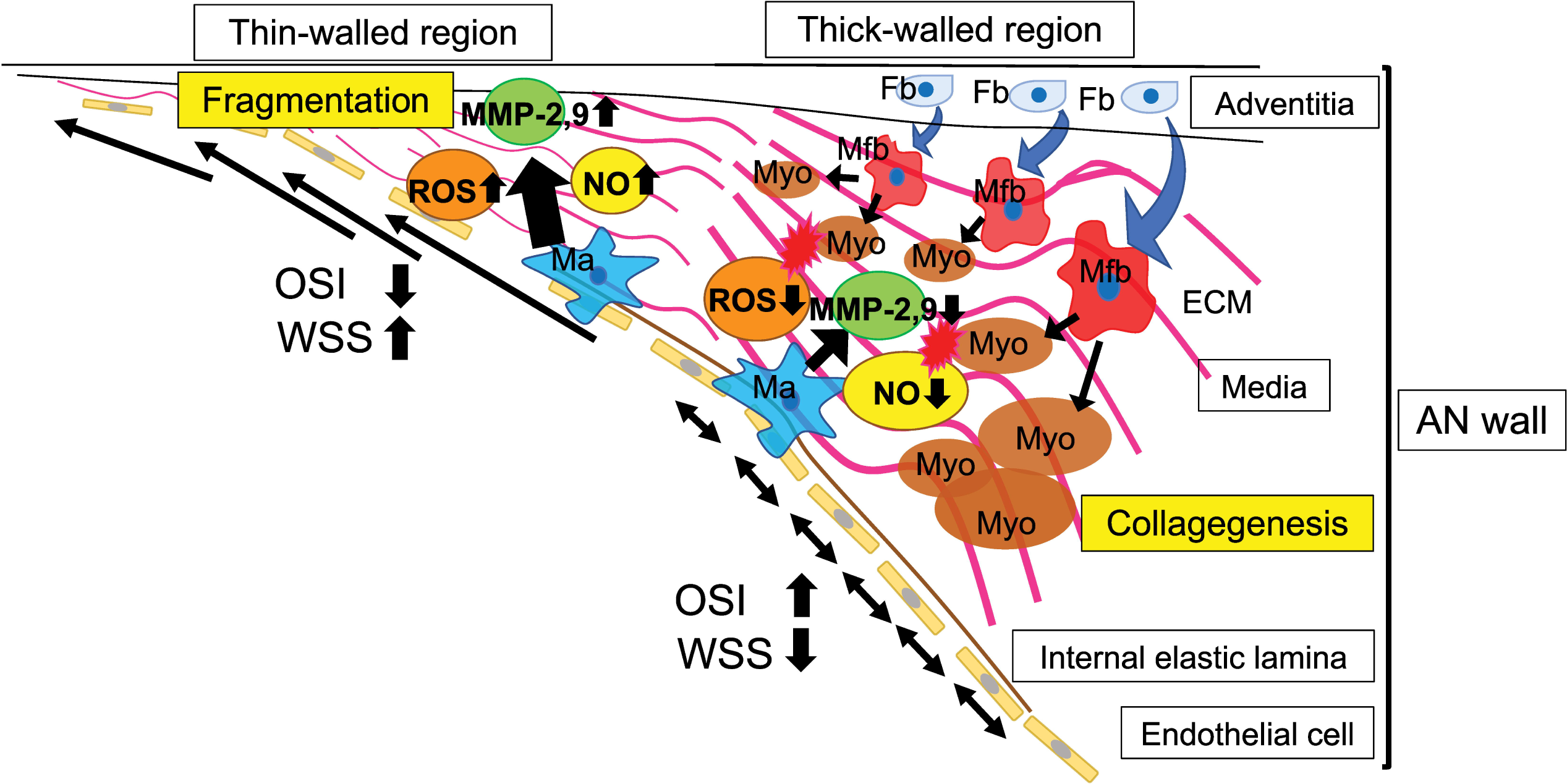
The possible role of myoglobin in determining aneurysmal wall thickness. Hemodynamically, areas with high oscillations in wall shear stress vectors (high OSI, relatively low WSS) allow fibroblast differentiation into myofibroblasts. The myofibroblasts secrete myoglobin which inactivates NO and ROS, which in turn suppress MMP-2 and MMP-9, and reduce ECM degradation, resulting in relative wall thickening. In contrast, myoglobin is not produced in areas where the wall shear stress vector does not fluctuate (low OSI, relatively high WSS). In these areas, NO and ROS cause the activation of MMP-2 and MMP-9, leading to collagen fiber fragmentation and reduction, and resulting in wall thinning. AN wall, aneurysmal wall; ECM, extracellular matrix; Fb, fibroblast; Ma, macrophage; Mfb, myofibroblast; MMP-2,9, matrix metalloproteinase-2 and matrix metalloproteinase-9; Myo, myoglobin; NO, nitric oxide; OSI, oscillatory shear index; MMP, matrix metalloproteinase.

Hemodynamic research has demonstrated that biological changes in the aneurysmal wall are triggered by hemodynamic stress on the wall.^19, 36, 37^ In areas exposed to high wall shear stress (WSS), WSS vectors are less variable, oscillatory shear index (OSI) is low, and MMP-2 and MMP-9 are activated via NO and ROS derived from vascular endothelial cells and macrophages, resulting in ECM degradation, thinning of the media, and apoptosis of SMCs and fibroblasts.^36, 37^ In contrast, areas with low WSS are areas of relative stagnation of blood flow where OSI is high and inflammatory cell infiltration causes wall thickening changes.^19, 37^ The detailed mechanisms of wall thickening are still unknown; however, myoglobin, which was discovered in the thickened areas, may counteract wall-destructive substances such as MMP-2 and MMP-9 by scavenging NO and ROS, resulting in the wall thickening.

Meng et al. reported that SMCs migrate during aneurysmal wall enlargement. In this study, we demonstrated that at least half of the cells in the aneurysmal wall that were thought to be SMCs were possibly myofibroblasts, which are also α-SMA-positive.^37^ Myofibroblasts differentiate from fibroblasts, and it has also been reported that fibroblasts do not become α-SMA positive.^22, 38^ Fibroblasts are reported to originally reside in the adventitia of the arterial wall.^39, 40^ In the aneurysmal growth stage, the area where the wall shear stress vector fluctuates significantly during one cardiac cycle is considered to be the area where blood flow is relatively stagnant. This hemodynamic condition may allow fibroblasts in the adventitia to differentiate into myofibroblasts and produce myoglobin, which may prevent wall weakening and induce wall thickening (Fig. 6).

### Limitations

This study had some limitations. First, the sample size is small. Second, the molecular biological mechanisms related to wall thickening and thinning are likely to involve numerous complex factors, and this study only examined pre- and post-processes involving myoglobin. Third, this study was based only on the histopathological findings. Further investigation is required to clarify the role of myoglobin in aneurysmal walls, including its association with other mediators of wall thickening and thinning.

## Conclusion

This is the first study to detect myoglobin in cerebral aneurysmal walls and suggest that myoglobin is derived from myofibroblasts in the aneurysmal wall. We also found evidence that myoglobin may be involved in preventing aneurysmal wall thinning by scavenging iNOS and ROS. Further elucidation of this pathology may lead to a better understanding of the mechanisms of aneurysmal thinning and thickening. Aneurysms are believed to rupture at the thinning part of the wall. Understanding the process of wall thinning may lead to new treatment approaches for unruptured cerebral aneurysms.

## Data Availability

All data produced in the present study are available upon reasonable request to the authors

## Acknowledgements

We would like to thank Ms. Tatsuko Uno for assisting with the pathophysiological analysis and Editage (www.editage.com) for English language editing.

## Non-standard Abbreviations and Acronyms

α-SMA: α-smooth muscle actin
CFD: computational fluid dynamics
ECM: extracellular matrix
iNOS: inducible nitric oxide synthase
MMP: macrophage-derived matrix metalloproteinase
NO: nitric oxide
OSI: oscillatory shear stress
ROS: reactive oxygen species
SMC: smooth muscle cell
WSS: wall shear stress

---

**Supplementary Figure 1.** Flowchart of sample selection for proteome and immunohistochemical analysis

**Supplementary Figure 2.** Accuracy of co-localization ratios between the cell counts and the areas

Receiver operating characteristic curves in differentiating co-localization ratios of myoglobin/periostin cell count from those of myoglobin/SMA cell count (solid line) and co-localization ratios of myoglobin/periostin area from those of myoglobin/SMA area (dashed line). The values of area under the curve (AUC) are not significantly different between the cell count and the area (0.975 [95% CI 0.937 – 1] versus 0.867 [95% CI 0.734 - 0.999]), respectively; p = 0.0525).

