## Supplemental Figure for "Cerebral aneurysm walls contain myoglobin that is possibly produced by myofibroblasts and contributes to wall thickening"

Supplement figure 1 Flowchart of sample selection for proteome and immunohistochemical analysis

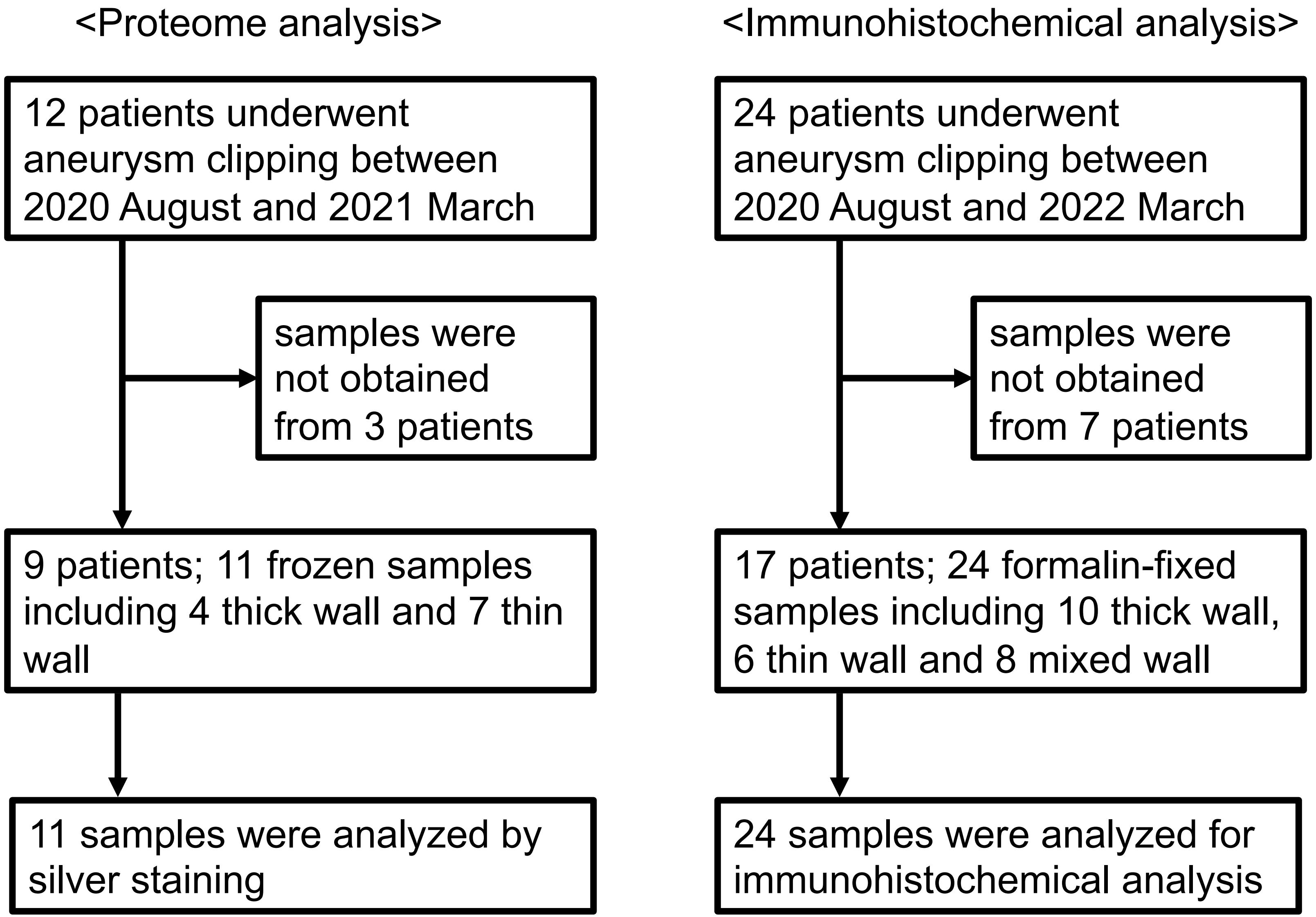

Supplement figure 2 Accuracy of co-localization ratios between the cell counts and the areas

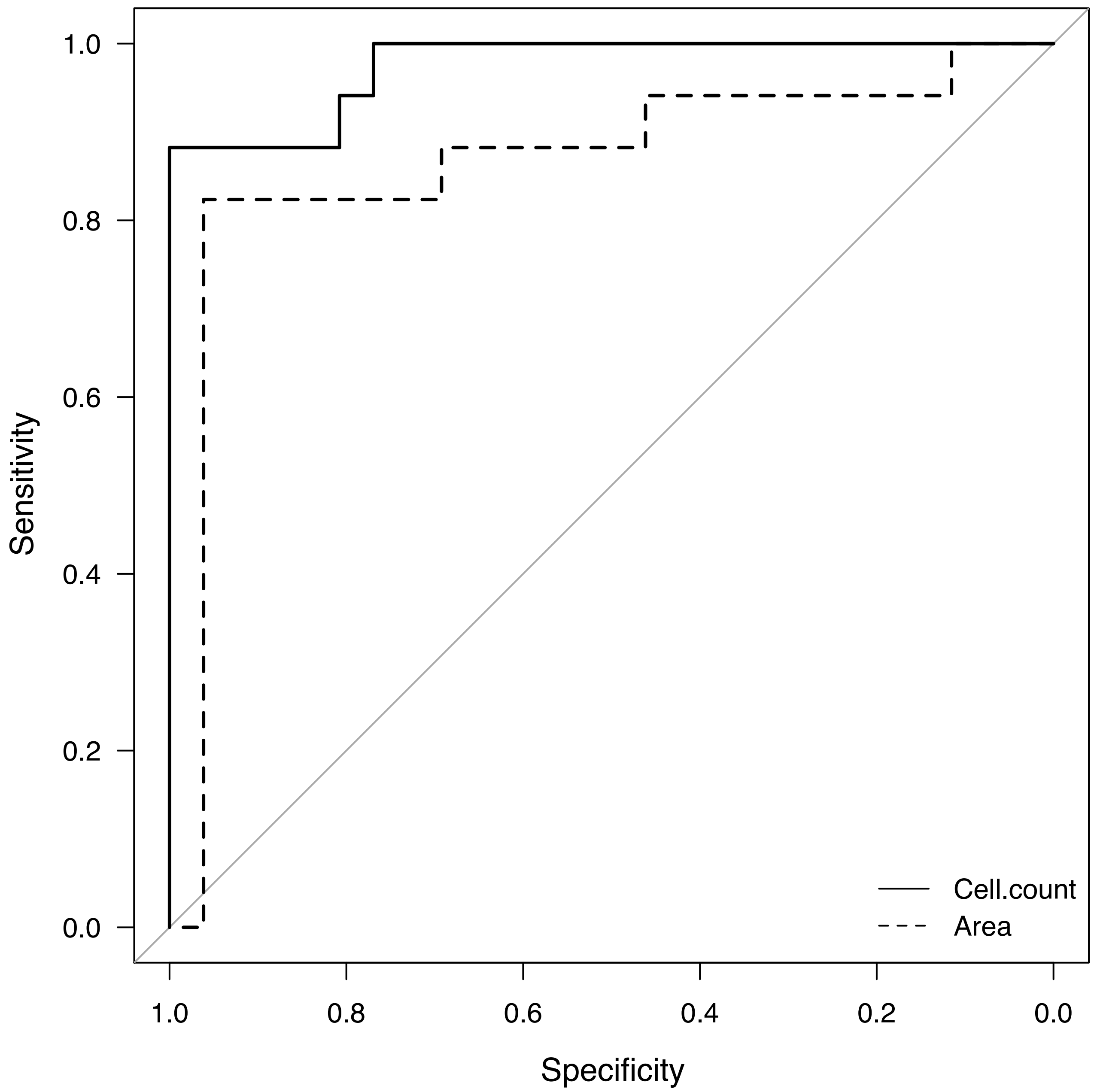


| | AUC | p value |
|---|---|---|
| Cell count | 0.975 | |
| Area | 0.867 | 0.0525 |
